# EARLY LIFE-COURSE PATTERNS OF REGISTRY-DEFINED SUBSEQUENT CANCERS AFTER HPV-RELATED MALIGNANCIES IN A U.S. POPULATION-BASED COHORT

**DOI:** 10.64898/2026.01.14.26344109

**Authors:** Jennifer M. Torres Del Valle, Claudia P. Amaya-Ardila, Souhail M. Malavé-Rivera

## Abstract

**Background:** Subsequent primary malignancies following human papillomavirus (HPV)–related cancers represent an important survivorship concern. However, evidence remains limited regarding sociodemographic and clinical factors associated with registry-defined subsequent cancers among children, adolescents, and young adults in U.S. population-based cohorts.

**Methods:** We conducted a retrospective population-based analysis of 1,326 individuals diagnosed with HPV-related cancers using Surveillance, Epidemiology, and End Results (SEER) data. Registry-defined subsequent cancer was operationalized as the occurrence of additional primary HPV-related malignancies according to SEER multiple primary rules. Multivariable logistic regression models estimated associations with sex, age group, area-level socioeconomic status (Yost Index quintiles), persistent poverty census tract status, and primary cancer site. Sex-stratified analyses by cancer site were performed.

**Results:** Registry-defined subsequent cancers were significantly associated with female sex and young adult age (20–29 years). Females had higher odds of subsequent cancer compared with males (OR = 1.06, 95% CI: 1.03–1.10), and individuals aged 20–29 years had higher odds than those aged 0–9 years (OR = 1.10, 95% CI: 1.05–1.16). Associations persisted after adjustment for socioeconomic indicators. No significant associations were observed with Yost Index quintiles or persistent poverty. Sex-stratified analyses showed higher odds of subsequent cancer for anal cancer among males and vulvar cancer among females relative to oropharyngeal cancer.

**Conclusions:** Sex and age are key determinants of registry-defined subsequent cancers following HPV-related malignancies, independent of area-level socioeconomic context. These findings support age- and sex-specific survivorship surveillance strategies across early life-course stages.

## Introduction

Human papillomavirus (HPV)–related cancers continue to pose a substantial burden in the United States despite advances in primary prevention, including vaccination. HPV is etiologically linked to malignancies across multiple anatomic sites, including the cervix, anus, vulva, vagina, penis, and oropharynx, with pronounced variation by age and sex (Saraiya et al., 2015; de Martel et al., 2017; Miller et al., 2022). Although the incidence of HPV-related cancers is lower in pediatric populations, diagnoses occurring in childhood, adolescence, and young adulthood represent a critical survivorship window due to long life expectancy and the potential for subsequent primary malignancies and long-term morbidity (Bleyer et al., 2006; Hudson et al., 2013; Oeffinger et al., 2006).

While a substantial body of literature has characterized incidence and survival patterns of HPV-associated cancers, comparatively fewer studies have examined subsequent cancer outcomes using large population-based datasets, particularly among children, adolescents, and young adults (AYA). This gap is especially relevant given evidence that AYA cancer survivors experience elevated risks of developing subsequent primary cancers and excess cancer-related mortality compared with the general population (Curtis et al., 2006; Travis et al., 2013; Sung et al., 2022). Survivorship challenges in this population are further compounded by transitions in care, fragmented follow-up, and inconsistent access to survivorship-informed healthcare services (Armenian et al., 2015; Bluethmann et al., 2016; Berkman et al., 2024).

Population-based cancer registries such as the Surveillance, Epidemiology, and End Results (SEER) Program provide a unique platform for evaluating long-term cancer outcomes at a national level (Howlader et al., 2020). However, SEER does not systematically capture clinically adjudicated recurrence, including local, regional, or metastatic relapses. Consequently, registry-defined subsequent primary malignancies have been widely adopted as a standardized and reproducible proxy for long-term cancer burden in survivorship research (Curtis et al., 2006; Travis et al., 2013; Wang et al., 2023). Although this approach does not capture clinical recurrence, it provides a consistent framework for population-based analyses when detailed clinical follow-up data are unavailable.

Emerging clinical and molecular evidence supports the plausibility of persistent HPV-related disease processes contributing to subsequent malignant events. For example, HPV integration status has been associated with residual or subsequent cervical disease following conization (Lin et al., 2025). Survivorship research in HPV-associated head and neck cancers has also highlighted long-term morbidity, quality-of-life impacts, and the need for coordinated survivorship care models (Osazuwa-Peters et al., 2019; Razzaghi et al., 2018). Together, these findings underscore the importance of population-based analyses examining subsequent cancer outcomes across early life-course stages. Accordingly, the objective of the present study was to evaluate sociodemographic and clinical factors associated with registry-defined subsequent cancers following HPV-related malignancies among children, adolescents, and young adults using SEER data.

## Methods

### Study Design and Population

We conducted a retrospective population-based cohort study using Surveillance, Epidemiology, and End Results (SEER) cancer registry data. The analytic cohort comprised 1,326 individuals diagnosed with HPV-related cancers across participating SEER regions in the United States. Individuals with both single and multiple primary cancers were included to enable assessment of registry-defined subsequent cancers.

### Database

The National Cancer Institute’s SEER program provides population-based cancer data on patient characteristics, tumor features, treatment, and outcomes. This study used a specialized SEER incidence dataset linking primary cancer sites with census tract–level socioeconomic status and rural–urban measures for cases diagnosed between 2010 and 2024 across 21 U.S. registries.

### Outcome Definition

The primary outcome was registry-defined subsequent cancer, operationalized as the occurrence of additional primary HPV-related malignancies following an initial cancer diagnosis according to SEER multiple primary rules. The outcome was analyzed as a binary variable (yes/no).

Because SEER does not capture clinically adjudicated recurrence, registry-defined subsequent primary malignancies serve as a standardized proxy for long-term cancer outcomes in population-based research (Wang et al., 2023).

### Independent Variables

Independent variables included sex, age group at diagnosis (0–9, 10–19, and 20–29 years), race, ethnicity, rural versus urban residence, persistent poverty census tract status, area-level socioeconomic status measured using the Yost Index (quintiles), and primary cancer site (oropharynx, anus, vulva, vagina, and penis).

### Statistical Analysis

We applied a multivariable analytic approach integrating demographic (sex, age), clinical (primary cancer site), and contextual factors (area-level socioeconomic status and persistent poverty) to examine associations with registry-defined subsequent cancers. Logistic regression models were used to estimate adjusted odds ratios and 95% confidence intervals. Sex-stratified analyses by cancer site were conducted to assess heterogeneity in associations.

### Ethics Statement

This study involved secondary analysis of de-identified data obtained from the Surveillance, Epidemiology, and End Results (SEER) Program. In accordance with federal regulations, this research was determined to be exempt from Institutional Review Board review.

## Results

Table 1 summarizes the sociodemographic and clinical characteristics of the study population stratified by registry-defined subsequent cancer status. Individuals with subsequent cancers were more likely to be female and to be diagnosed at ages 20–29 years compared with those without subsequent cancers (both p < .001). Differences by primary cancer site were also observed, with higher proportions of anal and vulvar cancers among individuals with subsequent cancers and a lower proportion of oropharyngeal cancers (p < .001). Distributions of race, ethnicity, rural– urban residence, persistent poverty census tract status, and Yost Index quintiles were similar between groups, with no statistically significant differences detected.

**Table 1.**
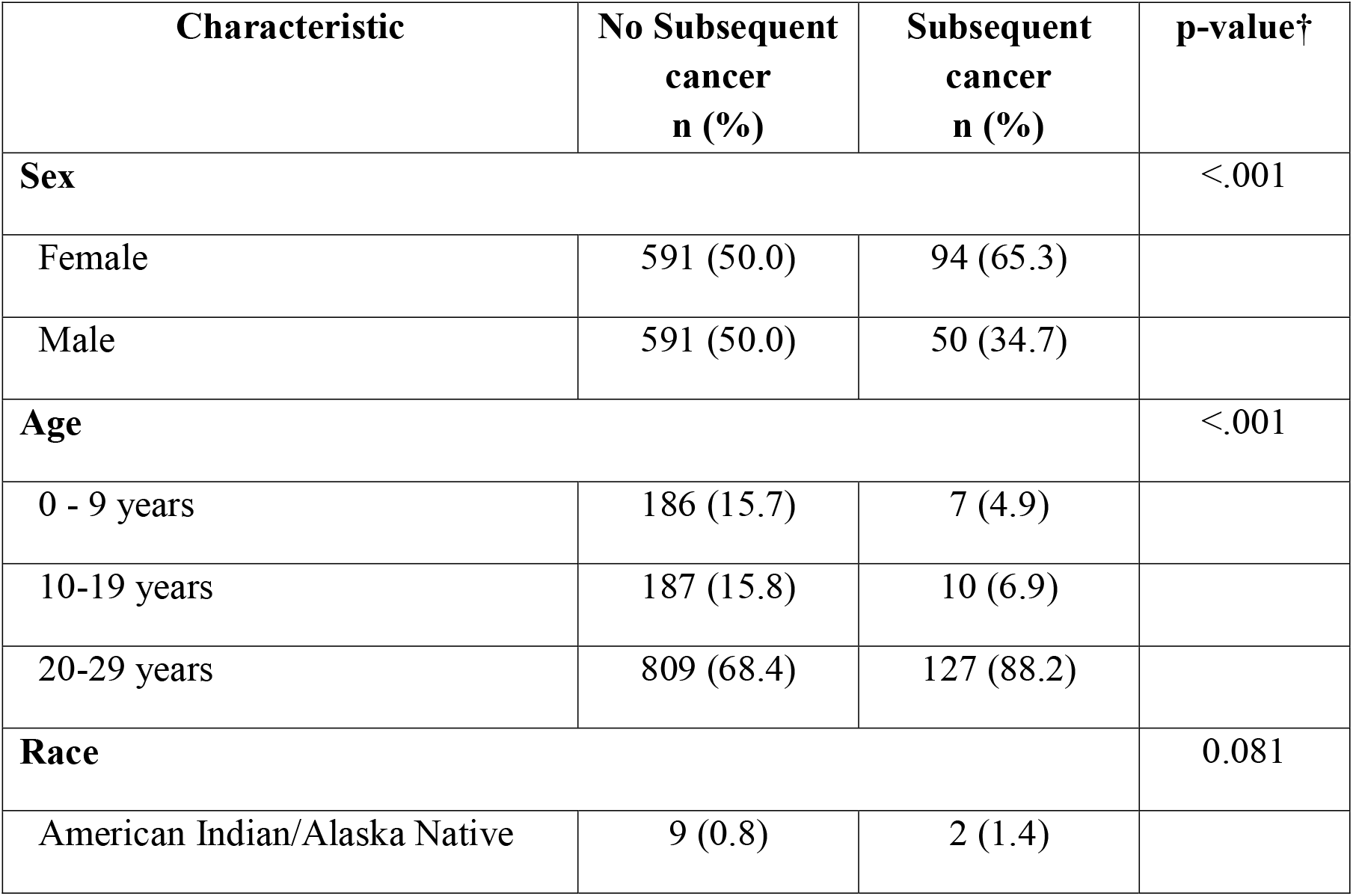
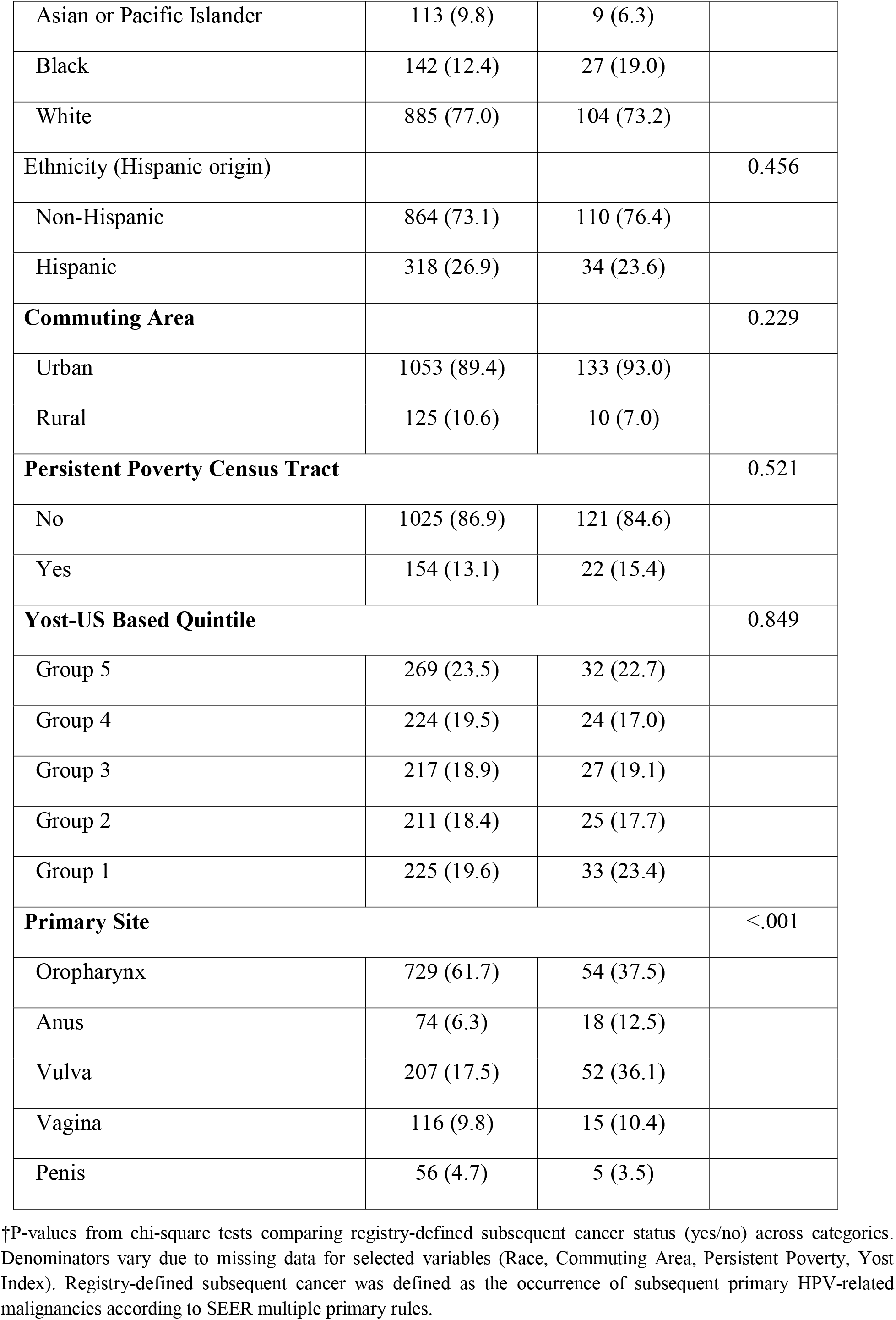
Sociodemographic and clinical characteristics of the study population.

Multivariable logistic regression results for registry-defined subsequent cancer are presented in Table 2. After adjustment, female sex was associated with higher odds of subsequent cancer compared with male sex (OR = 1.06, 95% CI: 1.03–1.10). Individuals aged 20–29 years had higher odds of subsequent cancer compared with those aged 0–9 years (OR = 1.10, 95% CI: 1.05–1.16), whereas no statistically significant association was observed for individuals aged 10– 19 years. Persistent poverty census tract status was not independently associated with subsequent cancer in adjusted models.

**Table 2.**
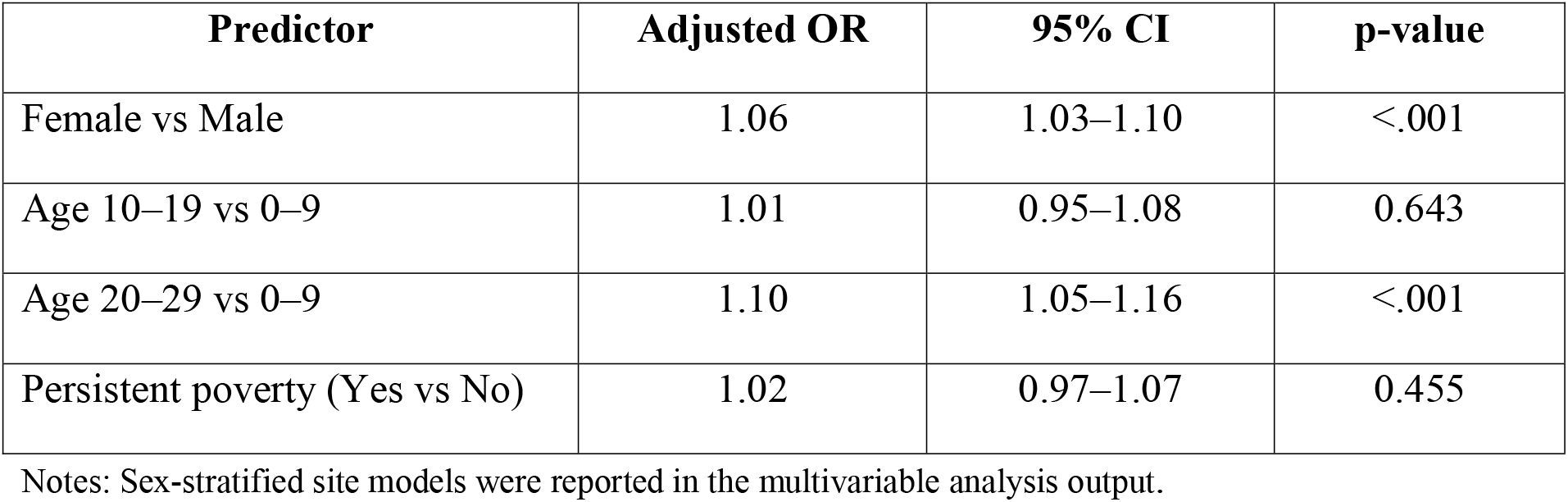
Logistic Regression Models for Registry-Defined Subsequent Cancers.

| Predictor | Adjusted OR | 95% CI | p-value |
| --- | --- | --- | --- |
| Female vs Male | 1.06 | 1.03–1.10 | <.001 |
| Age 10–19 vs 0–9 | 1.01 | 0.95–1.08 | 0.643 |
| Age 20–29 vs 0–9 | 1.10 | 1.05–1.16 | <.001 |
| Persistent poverty (Yes vs No) | 1.02 | 0.97–1.07 | 0.455 |

Notes: Sex-stratified site models were reported in the multivariable analysis output.

Sex-stratified multivariable models examining associations by primary cancer site are shown in Table 3. Among males, anal cancer was associated with higher odds of **registry-defined subsequent cancer** compared with oropharyngeal cancer (OR = 1.15, 95% CI: 1.08–1.23), whereas penile cancer was not significantly associated with subsequent cancer. Among females, vulvar cancer was associated with higher odds of subsequent cancer compared with oropharyngeal cancer (OR = 1.12, 95% CI: 1.06–1.19), while anal and vaginal cancers were not significantly associated with subsequent cancer.

**Table 3.**
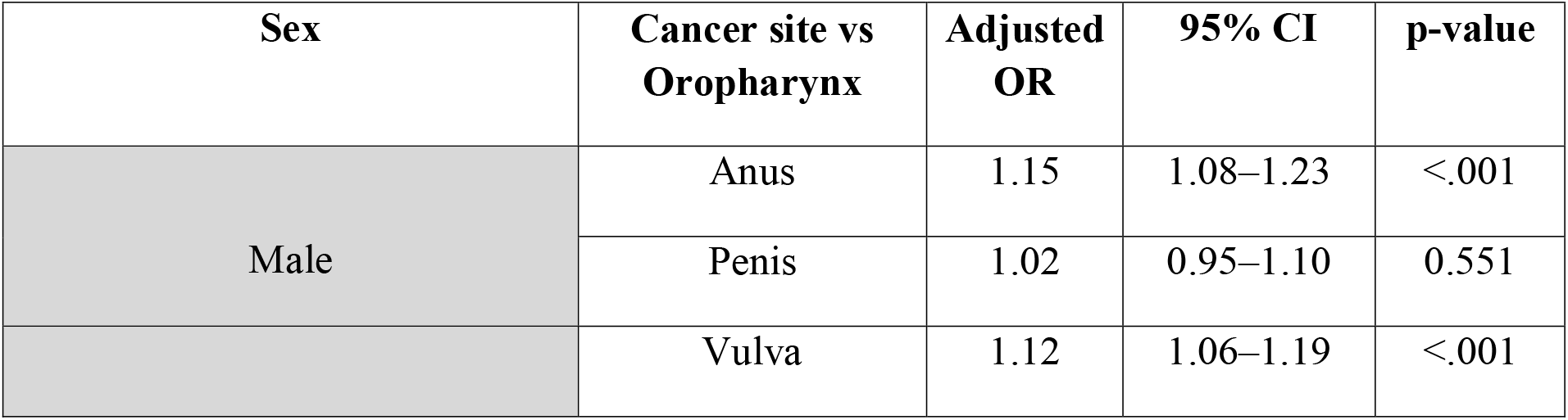
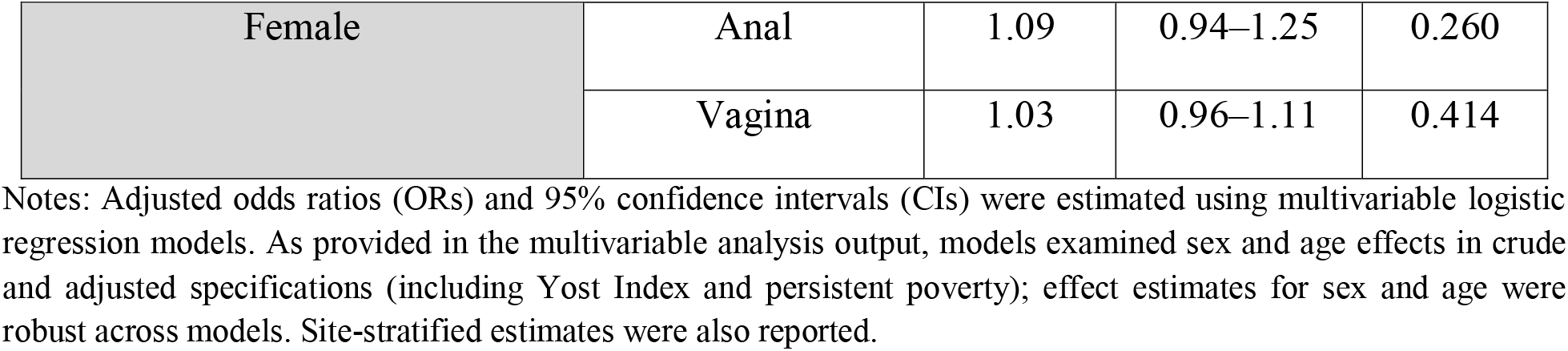
Sex-Stratified Cancer Site Models (Reference: Oropharynx)

**Figure 1.**
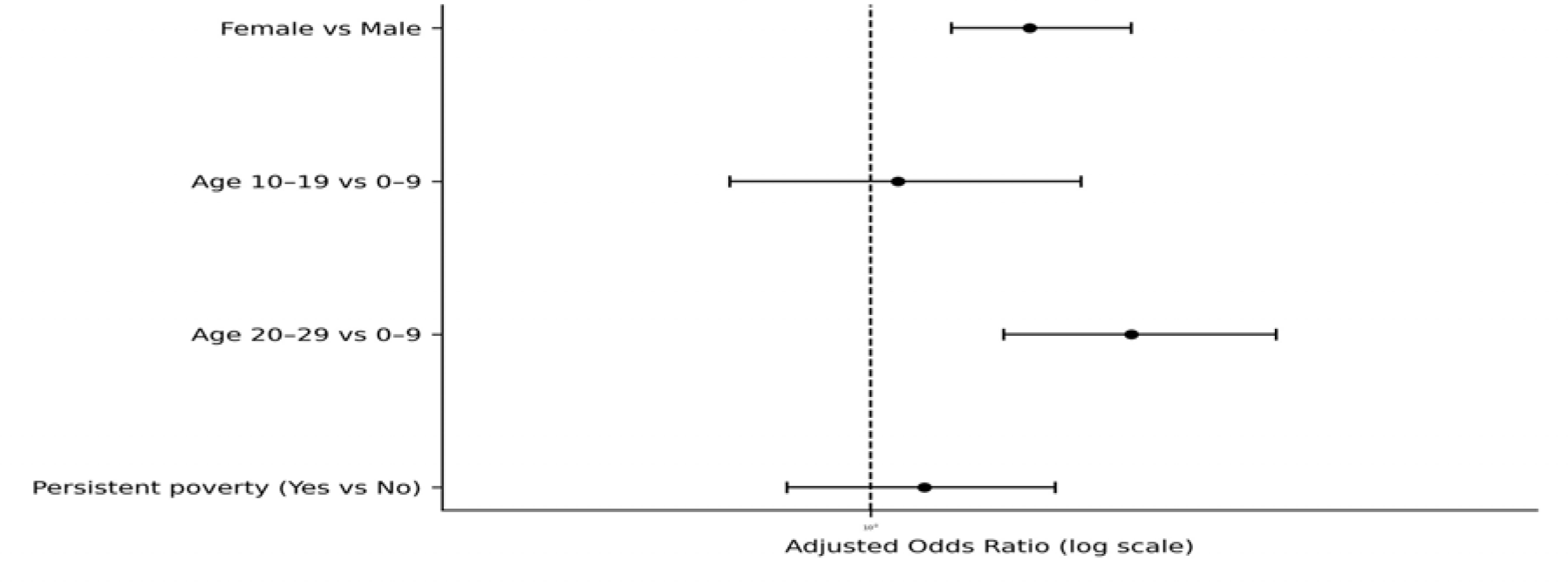
Adjusted odds ratios for HPV-related cancer recurrence. Estimates are derived from multivariable logistic regression models adjusted for sex, age group, primary cancer site, area-level socioeconomic status (Yost Index quintiles), and persistent poverty census tract. The vertical dashed line indicates the null value (OR = 1.0).

## Discussion

This study extends prior survivorship research by demonstrating that sex- and age-related differences in registry-defined subsequent cancers following HPV-related malignancies are detectable during early life-course stages. These findings align with national surveillance data showing heterogeneity in HPV-associated cancer burden by age and sex (Saraiya et al., 2015; Miller et al., 2022) and expand the existing literature by focusing on subsequent cancer outcomes among children, adolescents, and young adults.

The elevated odds of subsequent cancer observed among young adults are consistent with prior evidence indicating that adolescent and young adult cancer survivors experience increased risks of developing subsequent primary malignancies and excess mortality compared with the general population (Sung et al., 2022). Although transitions in care and long-term follow-up patterns were not directly measured in this study, survivorship frameworks suggest that disruptions during the transition from pediatric to adult oncology care may contribute to vulnerability during this life stage (Bluethmann et al., 2016; Berkman et al., 2024). These findings support the importance of age-specific surveillance strategies that extend beyond initial treatment completion.

Sex-specific differences by cancer site further highlight heterogeneity in survivorship trajectories. Higher odds of registry-defined subsequent cancer among males with anal cancer and females with vulvar cancer underscore the need for tailored surveillance approaches that consider anatomic site, sex, and developmental stage. Although registry-defined subsequent cancer does not capture clinically adjudicated relapses, this proxy provides valuable population-level insight into long-term cancer burden and survivorship risk.

The absence of independent associations between registry-defined subsequent cancer and area-level socioeconomic indicators should be interpreted cautiously. Area-level measures such as the Yost Index may not fully capture individual-level access to care, insurance coverage, or healthcare utilization patterns. Prior SEER-based research on HPV-differentiated nasopharyngeal carcinoma has demonstrated that socioeconomic contexts may differentially influence outcomes depending on HPV status and cancer site (Zhang et al., 2025), suggesting that access-related pathways not captured in registry-only analyses may contribute to observed disparities.

### Strengths and Limitations

Strengths of this study include the use of population-based SEER data, a focus on early life-course stages, and the integration of sociodemographic and clinical variables relevant to survivorship research. Limitations include the retrospective design and reliance on registry-defined subsequent primary malignancies as a proxy for clinically adjudicated outcomes. Additionally, socioeconomic status was assessed using an area-level measure that may not fully capture individual-level circumstances. Registry data also lacked individual-level healthcare utilization variables and detailed health insurance information, limiting the ability to directly examine access-to-care pathways, continuity of surveillance, and treatment-related factors that may influence subsequent cancer risk.

## Conclusions

In this population-based SEER cohort, sex and age were key predictors of registry-defined subsequent cancers following HPV-related malignancies among children, adolescents, and young adults. These findings support incorporating age- and sex-specific considerations into survivorship surveillance strategies across early life-course stages. Future studies linking registry data with clinical outcomes, healthcare utilization, and insurance information are needed to clarify access-related pathways and refine long-term follow-up care.

## Data Availability

The data used in this study are derived from the Surveillance, Epidemiology, and End Results (SEER) Program and are not publicly available. Access is subject to data use agreements and institutional approvals.

